# Dental students’ self-efficacy and intention to use clear communication techniques that support individuals with low health literacy

**DOI:** 10.1101/2023.07.16.23292737

**Authors:** Catherine Maybury, Alice M. Horowitz, Sharon R. Clough, Kerry M. Green, Min Qi Wang, Dushanka V. Kleinman, Cynthia Baur

## Abstract

**Objectives:** Clear and effective communication is critical to delivering quality dental care to all patients, especially those who have low health literacy. There is limited information about U.S. dental students’ exposure to communication techniques shown to improve patient understanding. Our primary objective was to assess rising fourth-year dental students’ knowledge, understanding, skills, self-efficacy, and behavioral intention to use communication techniques such as speaking slowly, using simple language and practicing the teach-back method. We also inquired about whether their dental curricula provided education about and evaluated them on the communication techniques.

**Methods:** This 2018 national cross-sectional study used a 34-item online survey to assess fourth-year dental students’ behavioral capability, self-efficacy and behavioral intention related to 17 communication techniques. The survey link was sent to 6,061 students; 242 finished it. Statistical analyses included descriptive statistics; ANOVA to examine associations between demographic variables and behavioral intention; and logistic regression to analyze associations between the predictor variables behavioral capability and self-efficacy and the dependent variable behavioral intention. The level of significance was set at p<0.05 for all analyses.

**Results:** Over 90 percent of students reported having the knowledge and skills to speak slowly, use simple language and teach-back, but they had lower self-efficacy and intention to use these techniques. They also were less knowledgeable, less confident and had lower intention to use other techniques. Students who reported higher self-efficacy were 9.2 times as likely to report higher behavioral intention to use the techniques than those who reported lower self-efficacy, 95% CI (4.10, 16.96), p<.01.

**Conclusions:** Our results indicate some dental students need additional education and training to increase their knowledge, skills, self-efficacy and behavioral intention related to plain language communication skills. It is possible that communication concepts are introduced in dental school but are not mastered at a level sufficient to be used in practice.

## Introduction

Clear and effective communication is critical to delivering quality dental care to all patients, especially those who have low individual health literacy (1,2). Low health literacy creates barriers for patients to access and understand health information and services, and it impedes their ability to take actions to protect and promote their health (3). Health literacy is influenced by both health system and individual level factors, and often there is a mismatch between the demands of the health care system and the skills and abilities of patients (3,4). Health care providers can decrease demands the health care system places on individuals by using communication techniques designed to increase patient understanding (4).

The *Healthy People 2030* definition of health literacy recognizes that organizations and health professionals have a responsibility to provide health information and services that support a patient’s ability to understand and use the information to care for their health (5). In addition, several of the *Healthy People 2030* Health Communication objectives are related to provider-patient communication (6). The new health literacy definition and the provider-patient communication objectives support and reinforce the need for clear communication between providers and patients.

Good dental provider-patient communication has several benefits (7). Effective provider communication can help patients understand health information, their oral health status and treatment options, and support their ability to make informed decisions and take appropriate actions to maintain their health (3,8,9). Good communication skills allow the provider to ask questions, listen, respond to patient concerns, demonstrate empathy, and provide health information and guidance at a level the patient can understand (9–11). Importantly, effective dentist-patient communication is associated with patients having regular dental care and improved adherence to treatment plans (10,11); decreased patient anxiety (12); and higher levels of oral health literacy (13,14).

We want to increase patients’ oral health literacy because those with lower levels of oral health literacy are more likely to have lower levels of oral health knowledge and less likely to use preventive regimens than those with adequate oral health literacy (3,10–12,15,16). They are less likely to ask questions of their dental provider or to ask their provider to explain information they do not understand (15,17,18). If individuals do not ask questions when they do not understand information or instructions, providers are less likely to know there is a problem. When dentists communicate clearly and confirm patient understanding, they increase the likelihood their patients understand health information, develop appropriate self-care routines and adhere to treatment plans (10,19), which can lead to better oral health outcomes.

Unfortunately, research indicates that many providers, including dental providers, do not routinely use clear communication techniques shown to improve patient understanding (20–24). In the U.S., dental and public health organizations have recommended changes to dental school curricula to prepare students to effectively communicate with people with all levels of health literacy (14,25–30). In addition, the U.S. Commission on Dental Accreditation (CODA) developed competencies for graduating dentists that included communication and interpersonal skills because “clear, accurate and effective communication is an essential skill for dental practice” (1).^(p454)^ These competences have been in place since 2009, but there is limited information about the approaches dental schools have taken to incorporate the communications and interpersonal skills competency into their curricula (15,16,31–36). Thus, the American Dental Association (ADA) Council on Advocacy for Access and Prevention (CAAP) and its National Advisory Committee on Health Literacy in Dentistry (NACHLD), conducted a national survey of rising fourth-year dental students in 2018 to explore their exposure to, confidence in using and intention to use communication techniques shown to improve patient understanding, which supports patients taking actions to protect and promote their oral health.

## Methods

### Study Design and Instrument Development

This cross-sectional study used a 34-item online survey to assess students’ behavioral capability, self-efficacy and behavioral intention related to 17 communication techniques. We adapted our survey from a national survey of general dentists that assessed routine use of 18 communication techniques shown to promote patient understanding (20) and from studies of Maryland health care providers’ use of these communication techniques (21–24). The communication techniques are described in the “Measures Section.”

The University of Maryland drafted the survey for the ADA in the spring 2017. Seven fourth-year dental students reviewed it and provided feedback on question clarity, survey flow and the time required to complete the survey. The survey was revised based on feedback from the students, the CAAP, and the ADA/CODA/ADEA Joint Advisory Committee on Dental Education Information (ACDEI). ADA CAAP staff then created an online survey using Qualtrics XM (37) and ADA’s Health Policy Institute (HPI) staff tested the survey to ensure it worked as we designed it. The ADA HPI administered the survey, and we (the University of Maryland) analyzed the data.

The ADA Council on Advocacy for Access and Prevention (CAAP) and its National Advisory Committee on Health Literacy in Dentistry (NACHLD) approved this study. In addition, we submitted the study to the University of Maryland Institutional Review Board (IRB) for ethical approval to analyze the data. The University of Maryland IRB exempted this study from review because it was considered secondary data analysis, e.g. the University of Maryland did not collect the data but we analyzed a de-identified data file the ADA provided us.

The ADA sent all fourth-year dental students an email that explained the purpose of the study, stated that respondents’ responses were confidential and they could not be identified, and that the survey was voluntary and they could stop at any time. The online survey used implied consent, which is the ADA’s protocol for online surveys. The email stated that by clicking on the ‘Start Survey’ button the students consented to participate in the study.

To increase participation, CAAP sent an email to all dental school deans asking them to encourage students to complete the survey. They sent a letter to American Dental Education Association (ADEA) senior management requesting they encourage faculty to support the survey. CAAP also offered a raffle of five $100 Amazon gift cards to students who completed the survey and elected to be in a drawing.

### Sample

The American Student Dental Association (ASDA) provided HPI with fourth-year dental student email addresses (n=6,123). ASDA reported that 95% of U.S. dental school students were members of their organization (ASDA staff, oral communication, April 2018). CAAP staff sent students an email that had a link to the confidential survey in July 2018. The survey was open for eight weeks; three follow-up reminders were sent. HPI staff provided us a de-identified data file for analysis in October 2018.

### Measures

To ground our research in behavioral theory, we based our study measures on Social Cognitive Theory (SCT) and Theory of Planned Behavior (TPB) constructs because of their predictive validity (38–41). We measured two SCT constructs – Behavioral Capability and Self-Efficacy and one TPB construct – Behavioral Intention (38–41). The survey asked students about their knowledge/understanding, skills, self-efficacy and behavioral intention related to each of 17 communication techniques (Figure 1).

**Figure 1.**
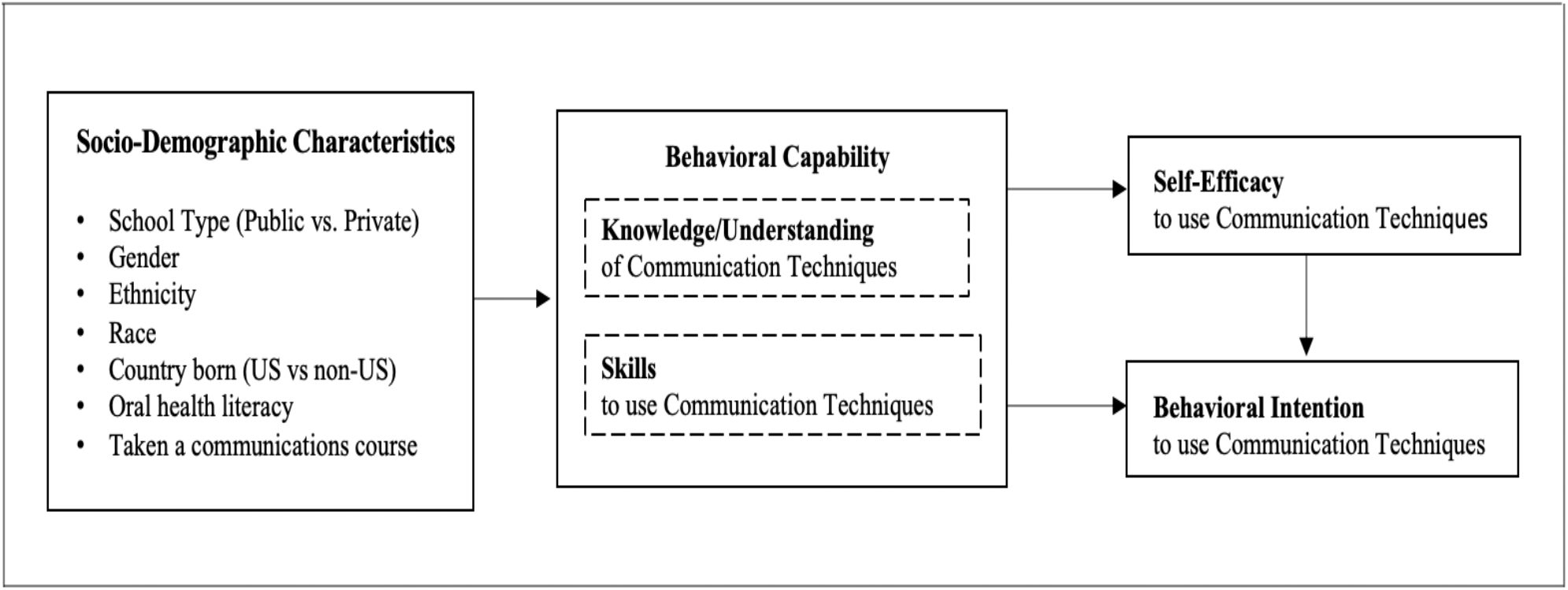
Conceptual model of dental students’ behavioral capability, self-efficacy and behavioral intention to use selected communication techniques with patients.

#### Predictor Variables

*Behavioral Capability* is the knowledge/understanding and skills to use 17 communication techniques (38). The predictor variables, *knowledge/understanding* of the communication techniques and *skills* to use the techniques, had response options of ‘yes’ = 1 or ‘no’ = 0, with 17 items each. The knowledge/understanding and skills items (n=34) were summed to create the behavioral capability index; values ranged from 0-34.

*Self-efficacy* is the confidence to use the techniques (39). Self-efficacy had response options of ‘not at all confident’ = 1, ‘somewhat confident’ = 2 and ‘extremely confident’ = 3. The items (n=17) were summed to create the self-efficacy index; values ranged from 17-51.

#### Dependent Variable

*Behavioral intention* is the primary outcome variable and it indicates the likelihood of performing a behavior (40,41) – using the communication techniques with patients after graduating from dental school. It had response options of ‘not at all likely’ = 1, ‘somewhat likely’ = 2 and ‘extremely likely’ = 3. The items (n=17) were summed to create the behavioral intention index; values ranged from 17-51. Behavioral intention is an indication of an individual’s likelihood of performing a behavior and it is considered the most proximate predictor of behavior (40,41). We assessed behavioral intention instead of ‘actual’ behavior because dental students are in school and the behaviors they perform are directed by the dental program and we wanted to understand behaviors students intend to adopt when they practice in a clinical setting outside of the dental school.

#### Demographic and Communication Skills Education Measures

The survey asked students if they had received communication skills education and training in their dental program (‘yes’ or ‘no’), if they had been evaluated on the use of the communication techniques (‘yes’ or ‘no’) and where they were evaluated (‘class only’, ‘clinic only’, or ‘class and clinic’). It also asked about the types of interpersonal skills taught in their dental school, educational methods and their greatest challenge communicating with patients who have difficulty understanding them. Demographic variables included their type of school (‘public’ or ‘private’), the country they were born in (‘U.S.’ or Non-U.S.’), gender, ethnicity, race, if they had ever taken a communications course (‘yes’ or ‘no’), and if they had heard the term ‘oral health literacy’ (‘yes’ or ‘no’).

### Power analysis, sample and data collection

We conducted an a priori power analysis using G*Power version 3.1.9 to determine the minimum sample size needed to adequately power our study. There was no guidance on effect size in the literature for oral health studies similar to the proposed research. Thus, we ran the sample calculation multiple times using small and medium effects based on Cohen’s definition of effect sizes (42). After consultation with a coauthor with statistical expertise, we decided to use a small effect size value of 0.2. According to G*Power, our study was considered adequately powered with a sample size of 117 (n=117). The following parameters were used in the power calculation: Test Family (F test), effect size (Cohen *d* = 0.2) (42); power (0.95); p-value (0.05) and 7 predictors.

### Statistical analysis

We analyzed the data using IBM SPSS Statistics version 25 (Armonk, N.Y.). The analyses included distributions (frequencies and percentages) and ANOVA to examine the associations between all demographic variables and the mean number of communication techniques. In the ANOVA, we used selected socio-demographic variables as predictor variables and the mean number of communication techniques for self-efficacy and behavioral intention as the dependent variables. We used logistic regression to analyze associations between predictor variables (behavioral capability and self-efficacy) and the dependent variable (behavioral intention). For the regression analyses we categorized the scores of each of the indices into three approximately equal groups to represent low, medium and high scores, following the approach used by the national and state studies^14–18^. The low and medium groups were combined to be the reference group in the logistic regression analysis comparing scores of the low/medium group to the group with high scores.

We ran four regression models to examine whether a) higher behavioral capability predicted higher self-efficacy, b) higher self-efficacy predicted higher behavioral intention, c) higher behavioral capability predicted higher behavioral intention, and d) when both higher behavioral capability and higher self-efficacy are included in the model, do both continue to independently predict behavioral intention (Figure 1). The models included socio-demographic variables that were significant in the bivariate analysis and were removed if they were no longer significant in adjusted models. We also compared characteristics of respondents to their cohort to assess response bias. The level of significance was set at p<0.05 for all analyses.

## Results

### Sample description

HPI staff sent the survey to 6,123 email addresses; 61 were invalid and one was a duplicate. In our sample of 6,061 students, 344 started the survey and 242 submitted it for a response rate of 4.0%. Respondents were from 55 of the 66 U.S. dental schools. The majority identified as female (61.2%), white (61.6%), attended a public institution (51.2%), were born in the U.S. (73.1%), and had heard of health literacy or oral health literacy (91.7%) (Table 1). Respondents differed significantly from fourth-year dental students generally in that they were more likely to be female (61.2% vs 48.8%, p<.01) and white (61.6% vs. 54.4%, p=.03) (43).

**Table 1.** Demographic characteristics of participating fourth-year dental students.

| Participant Characteristics (n=242) | n (%) |
| --- | --- |
| Type of Dental School |  |
| Public | 124 (51.2%) |
| Private | 118 (48.8%) |
| **Country of Birth** |  |
| US | 177 (73.1%) |
| Not US | 65 (26.9%) |
| **Gender** |  |
| Female | 148 (61.2%) |
| Male | 94 (38.8%) |
| **Ethnicity Hispanic or Latino<sup>†</sup>** |  |
| No | 219 (90.5%) |
| Yes | 22 (9.1%) |
| **Race** |  |
| White | 149 (61.6%) |
| Asian | 65 (26.9%) |
| Black/African American | 10 (4.1%) |
| More than one race selected | 9 (3.7%) |
| Race unknown | 8 (3.3%) |
| Native Hawaiian/Pacific Islander | 1 (0.4%) |
| **Ever Taken a Communications Course** |  |
| Yes | 143 (59.1%) |
| No | 99 (40.9%) |
| **Have Heard of Oral Health Literacy** |  |
| Yes | 222 (91.7%) |
| No | 15 (6.2%) |
| Don't know/Not sure | 5 (2.1%) |
| **Plans after graduation<sup>†</sup>** |  |
| Practice as an associate & be employed by another dentist | 79 (32.6%) |
| Practice as an associate in a corporate practice | 19 (7.9%) |
| Practice as a self-employed dentist | 5 (2.1%) |
| Enroll in a residency program in a dental specialty | 47 (19.4%) |
| Enter General Practice Residency/Advanced education general dentistry program | 56 (23.1%) |
| Practice in a community clinic/Federally Qualified Health Center | 21 (8.7%) |
| Practice in an Indian Health Service clinic | 4 (1.7%) |
| Unsure | 10 (4.1%) |
| Plan to pursue an MPH degree/MPH residency | 0 (0.0%) |

<sup>†</sup>Does not total to 242 due to missing data

### Communication skills education

Eighty-six percent responded they had received education and training related to the 17 communication techniques, but only 66.1% reported being evaluated on the techniques. Fifty-nine percent indicated they had ever taken a communications course; of these, 21.9% reported taking a communications course in dental school and 26.0% reported having lectures on communications that were integrated in courses in the dental school curricula. For educational methods used in their program, 84.0% reported they had lectures on provider-patient communication, 69.8% reported that written scripts were used to educate patients and 64.0% indicated standardized patients/actors were used for training (Table 2). When asked about their greatest challenge communicating with patients who have trouble understanding them, the most frequent responses were ‘I do not speak the patient’s language and I do not know what to do’ (20.7%) and ‘I need more experience explaining information in plain language’ (17.8%).

**Table 2.** Fourth-year dental students’ responses to questions about communication skills courses, evaluation and suggestions for communication skills training in dental school.

| **Education related to communication skills (n=242)** | **n (%)** |
| --- | --- |
| **Did your dental program provide communication skills education?** |  |
| Yes | 208 (86.0%) |
| No | 34 (14.0%) |
| **Did your dental program evaluate you on the 17 communication techniques?** |  |
| Yes | 160 (66.1%) |
| No | 82 (33.9%) |
| **Did your dental program evaluate you on the 17 communication techniques??** |  |
| Classroom & clinic | 116 (47.9%) |
| Classroom only | 30 (12.4%) |
| Clinic only | 14 (5.8%) |
| Question skipped because response of ‘No’ on previous question | 82 (33.9%) |
| **Should a communications skills course be taught in dental school?** |  |
| Yes | 125 (51.7%) |
| No/Don’t Know | 18 (7.5%) |
| No response | 99 (40.9%) |
| **In what year would a communication skills course be most beneficial?** |  |
| Second year | 82 (33.9%) |
| Third year | 75 (31.0%) |
| Fourth year | 30 (12.4%) |
| First year | 28 (11.6%) |
| **Where should a communication skills course be taught?** |  |
| Both lecture and clinic | 95 (39.3%) |
| Lecture only | 23 (9.5%) |
| Clinic only | 6 (2.5%) |
| **What types of interpersonal skills are taught in dental school? †** |  |
| Greet patients warmly | 213 (88.0%) |
| Encourage patients to ask questions | 208 (86.0%) |
| Consistently make eye contact | 206 (85.1%) |
| Sit rather than stand while talking with patients | 204 (84.3%) |
| Ask patients to explain their understanding of their dental problems | 179 (74.0%) |
| Give verbal or written information in multiple languages | 165 (68.2%) |
| Enlist help of patient’s family member/friend to promote understanding | 132 (54.5%) |
| Offer to help patients complete forms | 105 (43.4%) |
| **Which of these methods are used in your dental school? <sup>†</sup>** |  |
| Lectures on provider-patient communication | 203 (83.9%) |
| Interpreters or telephone translation for patients | 195 (80.6%) |
| Written scripts are used to educate patients | 169 (69.8%) |
| Standardized patients/actors are used for training | 155 (64.0%) |
| Patient education materials have been reviewed for readability and suitability | 147 (60.7%) |

<sup>†</sup>Yes/No response options for each item

### Descriptive results for communication techniques

Regarding having knowledge/understanding of each of the communication techniques, affirmative responses ranged from 98.3% to 55.8%; eight techniques had a response rate of 90% or higher. For skills to use the communication techniques, affirmative responses ranged from 96.3% to 59.9%; six techniques had a response of 90% or higher. For self-efficacy to use the communication techniques, responses of ‘extremely confident’ ranged from 79.8% to 52.9%; seven techniques had a response rate of at least 70%. For behavioral intention, responses of ‘extremely likely’ ranged from 83.1% to 37.6%, with five techniques having a response rate of 70% or higher (Table 3).

**Table 3.** Fourth-year dental students’ knowledge, skills, self-efficacy and behavioral intention to use seventeen communication techniques (n=242).

| Communication Techniques by<br>Domain and Item | Knowledge<br>to use<br>Technique | Skills<br>to use<br>Technique | Self-Efficacy<br>to use<br>Technique | Behavioral<br>Intention<br>to use<br>Technique |
| --- | --- | --- | --- | --- |
|  | 'Yes'<sup>a</sup> | 'Yes'<sup>a</sup> | 'Extremely<br>Confident'<sup>b</sup> | 'Extremely<br>Likely'<sup>c</sup> |
|  | n (%) | n (%) | n (%) | n (%) |
| **Interpersonal Communication** |  |  |  |  |
| Use simple language | 238 (98.3%) | 225 (93.0%) | 187 (77.3%) | 200 (82.6%) |
| Speak slowly | 235 (97.1%) | 228 (94.2%) | 190 (78.5%) | 189 (78.1%) |
| Present no more than 2 to 3 concepts at a time | 214 (88.4%) | 204 (84.3%) | 147 (60.7%) | 158 (65.3%) |
| Draw pictures or use printed illustrations | 211 (87.2%) | 199 (82.2%) | 162 (66.9%) | 161 (66.5%) |
| Ask if they would like family member/friend in the discussion | 180 (74.4%) | 196 (81.0%) | 151 (62.4%)<sup>†</sup> | 136 (56.2%) |
| **Teach-back Method** |  |  |  |  |
| Ask patients to repeat back information or instructions | 234 (96.7%) | 223 (92.1%) | 148 (61.2%) | 144 (59.5%) |
| Ask patients what they will do at home to follow instructions | 220 (90.9%) | 213 (88.0%) | 160 (66.1%) | 147 (60.7%) |
| **Assistance** |  |  |  |  |
| Read instructions out loud | 226 (93.4%) | 223 (92.1%) | 182 (75.2%) | 157 (64.9%) |
| Write or print out instructions | 219 (90.5%) | 219 (90.5%) | 191 (78.9%) | 177 (73.1%) |
| Follow-up by phone to check understanding and adherence | 203 (83.9%) | 206 (85.1%) | 156 (64.5%) | 145 (59.9%) |
| Underline key points on print materials | 202 (83.5%) | 205 (84.7%) | 183 (75.6%) | 155 (64.0%) |
| **Patient-friendly materials and aids** |  |  |  |  |
| Use models or x-rays to explain | 237 (97.9%) | 233 (96.3%) | 193 (79.8%) | 202 (83.1%) |
| Hand out printed materials | 216 (89.3%) | 215 (88.8%) | 191 (78.9%) | 180 (74.4%) |
| Use a video or DVD | 135 (55.8%) | 145 (59.9%) | 129 (53.3%) | 91 (37.6%) |
| **Patient-friendly Practice** |  |  |  |  |
| Use a translator or interpreter | 226 (93.4%) | 216 (89.3%) | 159 (65.7%) | 153 (63.2%) |
| Refer patients to Internet/other sources of information | 185 (76.4%) | 197 (81.4%) | 149 (61.6%) | 137 (56.6%) |
| Ask patients how they learn best | 156 (64.5%) | 176 (72.2%) | 128 (52.9%) | 128 (52.9%) |

<sup>a</sup> Response options were 'yes' or 'no'
<sup>b</sup> Response options were 'Extremely Confident', 'Somewhat Confident', 'Not at All Confident'
<sup>c</sup> Response options were 'Extremely Likely', 'Somewhat Likely', 'Not at All Likely'

### Bivariate and logistic regression analysis

In the bivariate analysis, none of the seven socio-demographic variables were significantly associated with self-efficacy (Table 4). Only country of birth was significantly associated with intention to use the techniques. Respondents born in the U.S. reported greater behavioral intention to use the techniques (mean 2.59) than those not born in the U.S. (mean 2.42), p=.025.

**Table 4.** Bivariate analysis of socio-demographic variables and self-efficacy and behavioral intention to use seventeen communication techniques (n=242).

| Characteristics | Sample Size<br>N (%)<sup>†</sup> | Self-Efficacy<br>to use 17<br>Communication Techniques |  | Behavioral Intention<br>to use 17<br>Communication Techniques |  |
| --- | --- | --- | --- | --- | --- |
|  |  | Mean<br>Score | Analysis of<br>Variance<br>(*P* value) | Mean<br>Score | Analysis of<br>Variance<br>(*P* value) |
| Type of School |  |  |  |  |  |
| Public | 124 (51.2%) | 2.63 | .65 | 2.57 | .22 |
| Private | 118 (48.8%) | 2.65 |  | 2.49 |  |
| Country of Birth |  |  |  |  |  |
| US | 177 (73.1%) | 2.65 | .18 | 2.58 | .03* |
| Not US | 65 (26.9%) | 2.58 |  | 2.42 |  |
| Gender |  |  |  |  |  |
| Female | 148 (61.2%) | 2.63 | .67 | 2.58 | .73 |
| Male | 94 (38.8%) | 2.65 |  | 2.53 |  |
| Ethnicity Hispanic or Latino<sup>†</sup> |  |  |  |  |  |
| No | 219 (90.5%) | 2.64 | .71 | 2.53 | .40 |
| Yes | 22 (9.1%) | 2.65 |  | 2.63 |  |
| Race |  |  |  |  |  |
| White | 149 (61.6%) | 2.65 | .55 | 2.54 | .94 |
| Non-white | 93 (38.4%) | 2.62 |  | 2.54 |  |
| Ever taken a communications course |  |  |  |  |  |
| Yes | 143 (59.1%) | 2.67 | .19 | 2.56 | .41 |
| No | 99 (40.9%) | 2.59 |  | 2.51 |  |
| Heard of Oral Health Literacy |  |  |  |  |  |
| Yes | 222 (91.7%) | 2.64 | .26 | 2.56 | .15 |
| No/Don't know | 20 (8.3%) | 2.55 |  | 2.39 |  |

\* $p < .05$
<sup>†</sup> Sample size less than 242 due to missing data.
<sup>‡</sup> The SE and BI indices ranged in value from 0-3.
SE= Self-efficacy; BI=Behavioral Intention

We ran four logistic regression models to examine the associations between our key variables (Table 5). Country of birth was nonsignificant in all models and removed from the final regression models. In Model 1, those who reported higher behavioral capability were 5.5 times as likely to have higher self-efficacy than those who reported lower behavioral capability, 95% CI (3.10, 9.80), p<.01. In Model 2, those who reported higher self-efficacy were 9.2 times as likely to have higher behavioral intention than those who reported lower self-efficacy, 95% CI (4.10, 16.96), p<.01. In Model 3, those who reported higher behavioral capability were 3.1 times as likely to have higher behavioral intention than those who reported lower behavioral capability, 95% CI (1.75, 5.34), p<.01. In Model 4, behavioral capability was not significant (p=0.10) when self-efficacy was entered into the model, and those who reported higher self-efficacy were 7.6 times as likely to have higher behavioral intention than those with lower self-efficacy, 95% CI (4.02, 14.55), p<.01.

**Table 5.** Logistic regression models predicting self-efficacy (Model 1) and behavioral intention (Models 2-4) to use seventeen communication techniques (n=242)

|  | Predictor Variable | Dependent Variable | *Odds Ratio* | *95% CI* | *P-value* |
| --- | --- | --- | --- | --- | --- |
| **Model 1** |  |  |  |  |  |
| Does higher behavioral capability predict higher self-efficacy to use 17 communication techniques? | Behavioral Capability | Self-Efficacy | 5.507 | (3.095, 9.799) | <.001 |
| **Model 2** |  |  |  |  |  |
| Does higher self-efficacy predict higher behavioral intention? | Self-Efficacy | Behavioral Intention | 9.202 | (4.992, 16.960) | <.001 |
| **Model 3** |  |  |  |  |  |
| Does higher behavioral capability predict higher behavioral intention? | Behavioral Capability | Behavioral Intention | 3.056 | (1.748, 5.342) | <.001 |
| **Model 4** |  |  |  |  |  |
| Do higher behavioral capability and higher self-efficacy predict higher behavioral intention? | Behavioral Capability | Behavioral Intention | 1.724 | (4.017, 14.550) | .100 |
|  | Self-Efficacy |  | 7.646 | (0.901, 3.297) | <.001 |
| Number of participants in low/medium (L/M) and High groups for each variable:<br>Behavioral Capability: L/M (n=94), High (n=148)<br>Self-Efficacy: L/M (n=140), High (n=102)<br>Behavioral Intention: L/M (n=119), High (n=123) |  |  |  |  |  |

## Discussion

Communication skills are recognized as an essential skill for delivering quality dental care (1,2) and communication and interpersonal skills are part of the competencies for graduating U.S. dentists (1). For dentists to have effective provider-patient communication skills, their undergraduate dental education must include communication skills education and training and foster practice using different scenarios so students are confident in using these techniques. Importantly, dental schools should evaluate students on these skills to ensure they meet communication/interpersonal skills competencies. Respondents’ self-reported knowledge, understanding, self-efficacy and intention to use the 17 communication techniques suggest some U.S. dental schools have implemented courses to support these competencies.

Yet, there is cause for concern about the emphasis placed on communication skills in some school curricula with only six in ten respondents indicating they had taken a communications course. Further, some students reported they needed more experience explaining information in plain language and others responded that they cannot provide information more simply than they already do. Student responses to questions about their self-efficacy and behavioral intention to use the communication techniques also indicate a need for greater emphasis on communication skills training. For self-efficacy, only seven techniques had a response rate over 70% for the ‘extremely confident’ response option. The highest percentage, 80%, was almost 20 points lower than the highest percentage for the knowledge and skills measures. More disconcerting was the low percentage of students who reported behavioral intention of ‘extremely likely’ to use the communication techniques with patients after graduation; only 5 of 17 techniques had a response rate higher than 70% (use simple language, speak slowly, write or print out instructions, use models or x-rays to explain and hand out printed materials). This may indicate students do not value the techniques or do not think they are important to their future practice. Also, one-third of students reported they had not been evaluated on the communication techniques. If students are not evaluated on a skill, they may deem it less important than skills they are evaluated on. Finally, with the widespread use of social media, dental schools have responsibility to teach their students methods to address inaccurate online health information and guide their patients to reliable information sources. On a positive note, respondents with higher levels of behavioral capability were more likely to have higher levels of self-efficacy and those with higher self-efficacy were more likely to have higher levels of behavioral intention. The theoretical underpinnings of our research support these findings (38–41).

Our findings are in line with results from the national and Maryland surveys of general dentists. The previous studies found dentists routinely used a low percentage of communication techniques, and our study found a low percentage of students who had high levels of self-efficacy and behavioral intention related to the techniques (20,21). In the previous studies the four communication techniques dentists reported using most frequently were the same four techniques associated with the highest level of behavioral intention in our survey – use simple language, use models or x-rays to explain, speak slowly, and hand out printed materials. A key difference between the previous surveys and this survey is the primary outcome measure. The previous surveys measured dentists’ routine use of communication techniques and our primary outcome measure was students’ behavioral intention to use these techniques because students are not yet practicing, and their practices are driven by the dental school practices. The outcome measures are different, but we think our findings provide insight into students’ behavioral intention, which is known to predict future behavior (40,41).

Dental schools can take several steps to place greater emphasis on communication skills in their program. Schools could conduct a health literacy environmental scan focused on communication skills (44,45); a scan could help schools assess faculty and staff education and training related to communication skills. A curricula review would assess the amount of time allocated to teaching communication skills and the context in which skills are taught. Additionally, schools could teach a standalone communications course that covers the depth and breadth of this important skill. Further, schools could adopt teaching methods that standardize patient interactions using written scripts for patient education and standardized patients/actors to allow students to practice provider-patient communications (46).

The current communication skills competencies are ten years old. Our findings suggest some dental schools are not adequately educating, training and evaluating students with regard to these competencies. CODA could take actions to require greater emphasis on communication skills in dental education. The CODA accreditation team could include members who have expertise in communication skills. These members could better elucidate current approaches to teaching these competencies and make recommendations to curricula and board examinations such that greater emphasis is placed on communication skills. Dental schools could hire communications or behavioral scientists to provide a voice for these skills in curricula design and provide accountability for implementing CODA communication skills competencies.

### Study limitations

The study had limitations. Our response rate was low. The timing of our survey (July to September) was unfortunate because that is when most fourth-year students begin seeing patients full time, which places increased demands on their time. Also, not all dental schools were represented in the study and the demographic characteristics of our participants differed from their nation-wide cohort, which limits our ability to generalize our findings. Further, respondents may have been more interested in communication techniques or attended a dental school that emphasized communication skills in their curriculum making them more likely to respond to the survey. If this were the case, our study results are likely to overreport the coverage of communication techniques in dental schools and the actual levels in the general fourth-year dental student population are lower than our results. Finally, our measures while based on established behavioral theory were exploratory in nature and had not been previously validated. In addition, the variables were highly skewed and this is likely due to using a 3-point scale. Future studies should explore using a 5- or 7-point scale to increase variability in responses.. However, the survey questions about communication techniques have been used in many studies of health care providers (7,20,21,23,24). Schools could explore using these constructs when revising or developing communication skills courses.

## Data Availability

All data produced in the present study are available upon reasonable request to the authors.

## Acknowledgements

This research was made possible by the Rima E. Rudd Dissertation Fellowship, awarded by the University of Maryland School of Public Health’s Horowitz Center for Health Literacy. Dr. Maybury received the fellowship in 2018.

Support in the form of services: The American Dental Association’s Council on Advocacy for Access and Prevention’s National Advisory Committee on Health Literacy in Dentistry, created the online version of the survey. The American Dental Association’s Council on Advocacy for Access and Prevention provided $500 for five $100 Amazon gift cards used for incentives.

